# The Efficacy of IL-6 Inhibitor Tocilizumab in Reducing Severe COVID-19 Mortality: A Systematic Review

**DOI:** 10.1101/2020.07.10.20150938

**Authors:** Avi Kaye, Robert Siegel

## Abstract

In the absence of highly effective antiviral therapies against SARS-CoV-2, it is crucial to counter the known pathophysiological causes of severe COVID-19. Evaluating the efficacy existing drugs may expedite the development of such therapeutics. Severe COVID-19 is largely the result of a dysregulated immune response characterized by lymphocytopenia, neutrophilia and critical hypercytokinemia, or “cytokine storm,” which is largely mediated by the cytokine interleukin-6 (IL-6). The IL-6 inhibitor tocilizumab (TCZ) could potentially suppress the effects of the pro-inflammatory cytokine and thereby lower mortality from the disease. This systematic analysis aimed to investigate and synthesize existing evidence for the efficacy of TCZ in reducing COVID-19 mortality. PubMed and SearchWorks searches were performed to locate clinical studies with primary data on TCZ treatment for severe COVID-19. Sixteen case-control studies comparing mortality between TCZ and standard of care (SOC) were identified for quantitative synthesis. Combined mortality for the TCZ-treated and SOC groups were 26.0% and 43.4% respectively. In all but one of the studies, the odds ratio of mortality from COVID-19 pointed towards lower fatality with TCZ versus the SOC. A combined random effects odds ratio calculation yielded an odds ratio of 0.453 (95% CI 0.376-0.547, p<0.001). Additionally, eighteen uncontrolled trials were identified for qualitative analysis producing a raw combined mortality rate of 16.0%. Important caveats to this research include the lack of prospective randomized control trials (RCTs) and the absence of data from the large COVATA study from the published literature. However, results from this systematic analysis of published research provide positive evidence for the potential efficacy of TCZ to treat severe COVID-19, validating the ethical basis and merit of ongoing randomized controlled clinical trials.

## Introduction

Coronavirus Disease 2019 (COVID-19) – caused by the novel Severe Acute Respiratory Syndrome Coronavirus 2 (SARS-CoV-2) – manifests in a broad range of disease severity. Roughly 85% of confirmed cases present as a mild respiratory illness defined as minor fatigue, low-grade fever and dry cough, 15% develop severe pneumonia requiring hospitalization and 5% become critical indicated by acute respiratory distress syndrome (ARDS), septic shock, and multi-organ failure resulting in ICU admission, mechanical ventilation, and death.^1^ The most significant known risk factors for COVID-19-related death are increasing age, chronic comorbidities including diabetes, cardiac disease, pulmonary and kidney dysfunction and male sex.^2^ A dysregulated immune response – characterized by decreased T-cell counts, increased inflammatory cytokines and extra-pulmonary systemic hyperinflammation syndrome – is principally responsible for inducing critical pulmonary failure observed in COVID-19 and largely driven by interleukin-6 (IL-6).^3^ This systematic review concerns the efficacy of an interleukin-6 inhibitor, tocilizumab (TCZ) in reducing severe COVID-19 mortality.

### COVID-19 Dysregulated Immune Response and the Role of IL-6

Severe COVID-19 is characterized by a dysregulated immune response to SARS-CoV-2 infection that is implicated in disease mortality even after viral load decreases.^4^ The immune dysregulation presents with two sequential and diametrically opposed reactions that both instigate symptom aggravation.^5^ The first pattern is impaired adaptive immune response with lymphocytopenia, which includes markedly reduced CD4+ and CD8+ T-cells, B-cells and natural killer (NK) cells. While T-cells are significantly decreased in all COVID-19 patients, reduction in B and NK cells are more affected in severe cases.^6,7^ Adaptive immune cell depletion impairs the body’s ability to clear the virus and mitigate inflammatory reactions. ^8^

The second deleterious response is an over-activation of the innate immune system. This pathogenic response is characterized by an increase in neutrophils and pro-inflammatory cytokines including IL-6, IL-1β, IL-2, IL-8, CCL3 and TNF-α. The rapidly increasing cytokine levels – also known as a “cytokine storm” – drives progression to septic shock, tissue damage and multiple organ failure (heart, liver, kidney, respiratory).^5^ The effects are instigated by excessive NF-κB pathway activation, alarmin release by damaged epithelial cells, neutrophil and macrophage infiltration, and alveolar damage by vessel permeability and alveolar wall thickening.^8^

Roughly three-quarters of patients in severe condition present with IL-6 mediated respiratory failure.^9^ IL-6 concentration is thus a reliable predictor of COVID-19 severity as it is significantly elevated in fatal cases.^10^ IL-6 has pleotropic effects including hematopoiesis, metabolic regulation, inflammation, autoimmunity and acute phase response.^11^ Some IL-6-dependent outcomes help stave off infections such as directing neutrophil migration to the infection site, increasing CD8+ T cell cytolytic capacity, and regulating antiviral thermostatic reactions. However, IL-6 is also implicated in viral infection disease progression as it leads to tissue permeability and edema, reduces IFN-γ production, drives anti-apoptotic molecules and promotes excessive neutrophil survival. The above adverse effects promote lethal inflammation and enable viral infiltration to distant organs.^12^ Furthermore, elevated serum IL-6 is associated with impaired cytotoxic activity of NK cells, thus weakening their virus-killing capacity.^13^ IL-6 is known to increase the rate of fibrotic clot formation, so it also may play a role in the thrombotic complications observed in COVID-19.^12^ The renin-angiotensin system, which controls blood pressure and electrolyte balance, is an additional important factor in IL-6 modulation and COVID-19 pathology. As virus binds ACE2, thus reducing its availability, there is an increase of angiotensin II in COVID-19 patients, creating a positive feedback loop that advances pro-inflammatory signaling. ^12^ The responses and physiological effects of IL-6 release are summarized in Figure 1.

**Figure 1:**
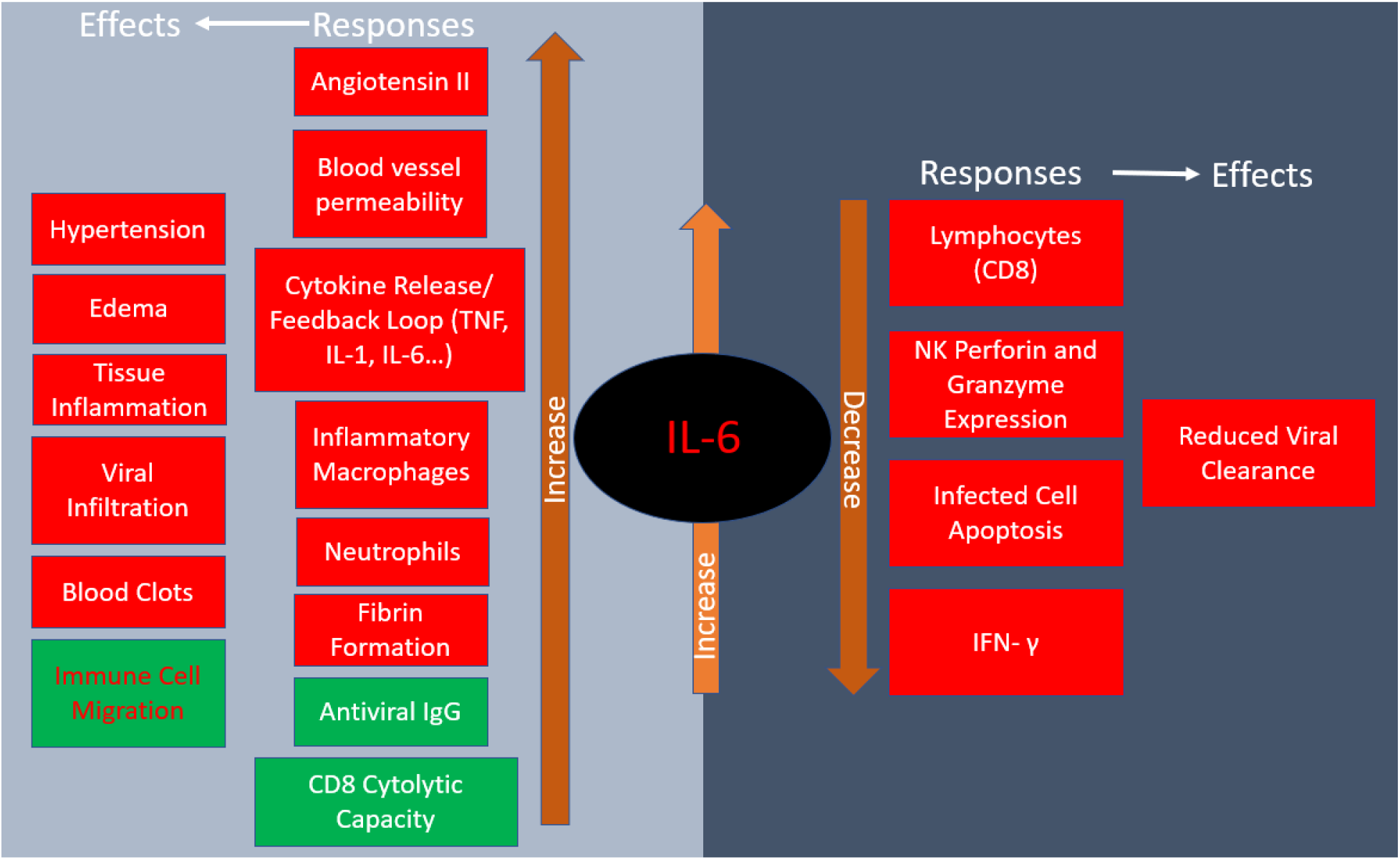
Responses to IL-6 release and their physiological effects. Red indicates negative consequences while green designates positive ones in COVID-19.

### COVID-19 Treatments Antivirals

Without a targeted drug for COVID-19, scientists and clinicians are attempting to rapidly find alternative treatments and solutions to combat the disease’s lethal immunological effects.^14^ One strategy is to utilize existing antiviral drugs with the expectation that they may exhibit similar effects against SARS-CoV-2. One of the more promising treatments is RNA-dependent RNA polymerase inhibitor remdesivir which is shown to reduce COVID-19 mortality but is less effective in severe cases.^15^ Other commonly used antivirals are hydroxychloroquine and chloroquine. Despite widespread utilization for COVID-19 patients, evidence is lacking for the clinical efficacy and prophylactic properties of hydroxychloroquine or chloroquine despite their *in vitro* antiviral and *in vivo* immunomodulatory properties.^16^

### Immune Suppressants

There are numerous investigations of existing immune suppressing and anti-cytokine interventions to counter the dysregulated, excessive immune response. After early evidence and recommendations against the use of corticosteroids to treat severe COVID-19,^17,18^ a large randomized evaluation of dexamethasone found that the drug significantly reduced 28-day mortality in patients included in the study (rate ratio 0.83; 95% CI 0.74-0.92; p<0.001). However, mortality rate reductions varied depending on baseline respiratory demands upon randomization as there was reduction for patients on mechanical ventilation and oxygen but not for patients without respiratory support.^19^ As of mid-August 2020, the WHO modified their recommendation against corticosteroids to include judicious administration under respiratory failure with ARDS.^20^ Because of the systemic effects of corticosteroids, more options for targeted immune regulation is warranted. Common targets for inhibition include IL-6, the IL-1 family (IL-1β and IL-18), TNF-α and IFN-γ cytokines and the JAK pathway.^8^

IL-6 is a particularly promising target due to its correlation with ARDS severity and mortality.^21^ IL-6 inhibitors are already successfully utilized for other cytokine storm syndromes such as adverse T cell therapy reactions and Still’s disease-associated hemophagocytic lymphohistiocytosis.^8^ In COVID-19, IL-6 inhibitors should be carefully administered with appropriate timing due to its control of viral replication.^22^

### IL-6 Inhibitor Tocilizumab

Tocilizumab (TCZ) [Actemra] is a recombinant monoclonal antibody with a humanized murine variable domain and a human IgG1 constant domain. TCZ binds to both membrane-bound and soluble IL-6 receptors, thus preventing IL-6 mediated signal transduction. The drug was initially developed to treat rheumatoid arthritis and now it is also approved for giant cell arteritis and similar autoimmune ailments. Its safety profile was analyzed in a phase III double-blind placebo-controlled trial and it is reportedly effective in treating other cases severe cytokine release syndrome such as chimeric antigen receptor T-cell immunotherapy.^11^

While TCZ is not yet approved for treatment of COVID-19, clinicians across the globe are utilizing the drug under emergency use authorization, including in the United States. One of the largest initial observational studies of TCZ evaluated 547 COVID-19 ICU patients in New Jersey comparing the survival rate of 134 individuals treated with standard of care (SOC) and TCZ with SOC controls finding a 46% and 56% mortality rate respectively and a 0.76 adjusted hazard ratio.^23^ However, there was insufficient statistical power to conclude clinical efficacy of TCZ with the clinical data and they only focused cases that already progressed to a critical stage leading to an inflated mortality rate for both groups.

Randomized controlled trials (RCT) are the gold standard for evaluating the clinical efficacy of a drug. The first analyzed RCT for TCZ presented negative results. Genentech (Roche) – the producer of TCZ – discontinued their 450-participant phase III trial COVACTA because it failed to meet the primary endpoint of improved clinical status after 4 weeks with TCZ versus the SOC.^24^ The disappointing outcome from COVACTA places serious doubt on the efficacy of TCZ against COVID-19. Additionally, another IL-6 inhibitor Sarilumab failed its Phase III RCT in the United States but still has some ongoing (NCT04322773, NCT04327388, NCT04412772).^25^ Other IL-6 inhibitors under investigation include siltuximab (SYLVANT, NCT04322188) and fingolimod (NCT04280588) Nonetheless, several phase III and phase II RCTs remain in progress into at least September (REMDACTA, NCT04409262; NCT04372186, NCT04356937) and healthcare providers are still currently administering TCZ globally for advanced COVID-19 cases. However, the logic of suppressing IL-6 remains convincing. Therefore, it is essential that the known impact of TCZ is analyzed in a systematic manner to ascertain whether its continuing use is ethical, even in RCTs. Since dexamethasone is a broadly acting immunosuppressant, it should be noted that the use of this drug in clinical trials may obscure the effects of more targeted immunomodulatory drugs like TCZ.

Existing systematic reviews investigate only case studies of TCZ treatment without controls.^26,27^ Therefore, this systematic review will synthesize the evidence from individual case-control studies and analyze uncontrolled trials to determine whether the drug is potentially effective at reducing severe COVID-19-related mortality, thus corroborating the logic for continuing RCTs to evaluate the potential use of IL-6 inhibitors.

### Systematic Review Methods and Statistics

Articles utilized for the systematic review were selected from a PubMed search on August 4, 2020. For the initial screening, the primary search terms were COVID-19 or SARS-CoV-2 and tocilizumab. Papers with primary data for a case-control study comparing mortality rate from severe COVID-19 between TCZ and standard of care (SOC) were included for data synthesis. Uncontrolled studies on severe COVID-19 mortality with TCZ were reviewed separately without data synthesis. Exclusion criteria included papers without primary data, case reports, reviews, protocols, and studies without mortality numbers available or potentially repeating patient data. An additional search was performed on SearchWorks to identify case-control studies not found in PubMed.

For each study included in the synthesis, the mortality rate for the TCZ and SOC group were calculated. In the controlled studies, the odds ratio (RR) of mortality from COVID-19 with TCZ versus the SOC was determined followed by the 95% confidence interval (CI) and p-value calculation. The data from the individual controlled studies were synthesized by a random effects meta-analysis calculation using MedCalc software. MedCalc was also used to perform a sample size calculation with an alpha of 0.01 and power of 90% to detect a difference between the total crude TCZ and SOC mortality rates.

The systematic review protocol was pre-registered with PRISMA and approved on June 22, 2020 (CRD42020193479).

## Results

A total of 314 articles were identified by the initial PubMed search and three additional case-control studies were found on a SearchWorks (Fig. 2).^28^ 38 articles were selected for full-text review yielding 16 uncontrolled studies for qualitative analysis and 18 case-control studies for both quantitative synthesis and qualitative analysis. The study characteristics for the controlled studies are summarized in Supplementary Table 1 while the uncontrolled studies are outlined in Supplementary Table 3.

**Figure 2:**
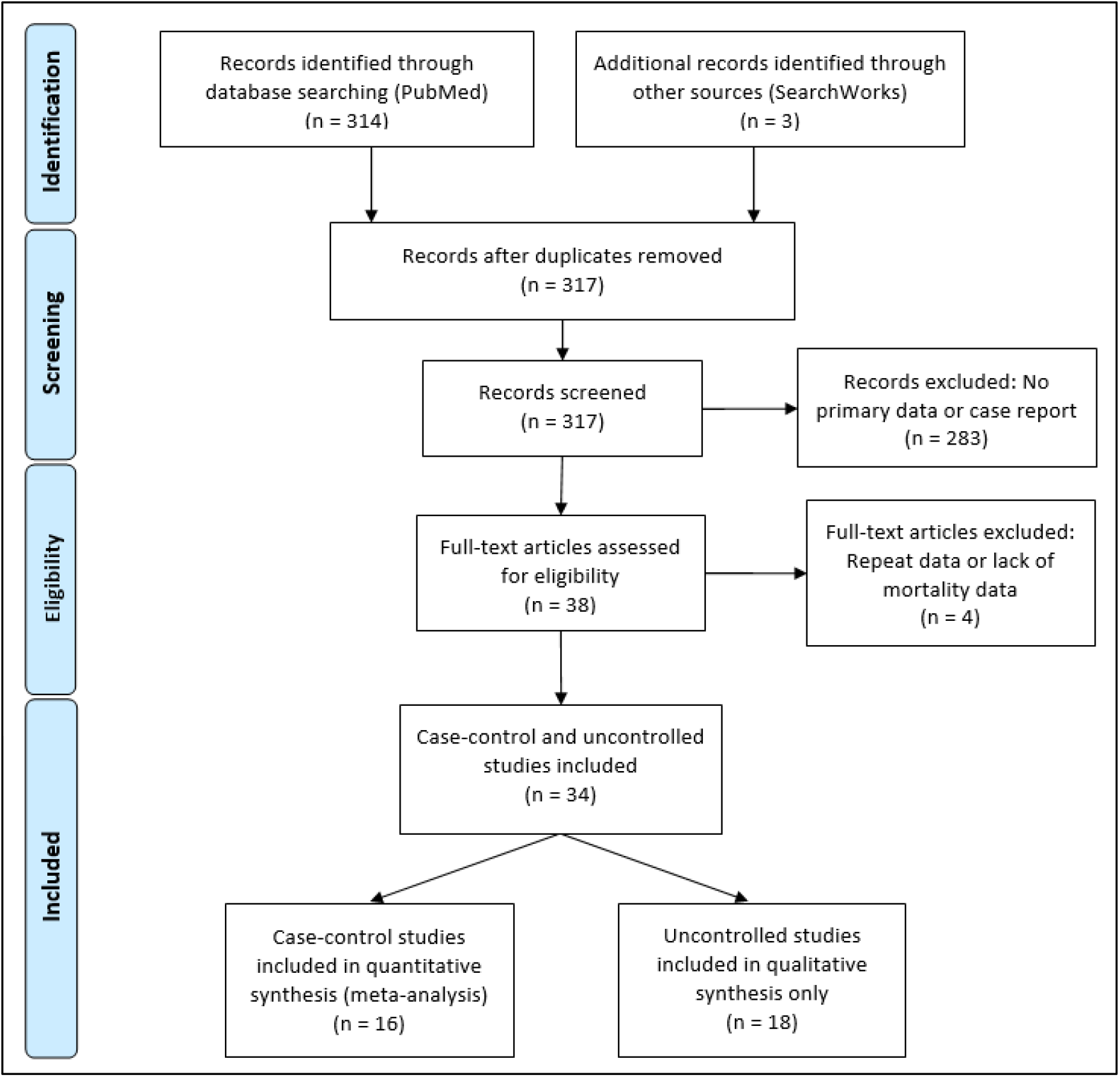
PRISMA flow diagram for systematic review article search.^28^

### Controlled Studies

The systematic review of sixteen controlled studies encompassed a total of 1008 TCZ-treated and 1537 SOC control patients (Table 1). 13 of the studies occurred in a single medical center while the remaining 3 aggregated data from multiple hospitals. The largest patient contributions to the analysis were from the multiple-hospital studies [Ip et al. and Guaraldi et al.] The baseline patient characteristic for all but one study [Guaraldi et al.] was severe COVID-19, generally qualified by oxygen supplementation needs. Ip et al., Eimer et al. and Potere et al. only analyzed patients who were already admitted into the ICU. 61% and 44.8% of both cases and controls in Rojas-Marte et al. and Roumier et al. respectively began the trial in critical condition. There was some variation in TCZ administration and SOC treatments, the most common being hydroxychloroquine – utilized in all but one study (Capra et al.) – and lopinavir/ritonavir. Length of observation ranged from 7 days to 30 days with endpoints of death or discharge. Mean age of participants in the treatment and control groups was 55.5 to 76.8 with no more than 6.1 years separating the two groups within one study.

**Table 1:** Quantitative synthesis of individual case-control study mortality data.

| Study | TCZ Mortality | Controls | Odds ratio | 95% CI | Weight %<br>(Random Effects) | p Value |
| --- | --- | --- | --- | --- | --- | --- |
| Klopfenstein et al.<sup>29</sup> | 4/20 (20%) | 12/25 (48%) | 0.271 | 0.0704-1.042 | 3.54 | 0.0575 |
| Campochiaro et al.<sup>30</sup> | 5/32 (15.6%) | 11/33 (33.3%) | 0.37 | 0.112-1.227 | 4.23 | 0.104 |
| Capra et al.<sup>31</sup> | 2/62 (3.2%) | 11/23 (47.8%) | 0.0364 | 0.00713-0.185 | 2.61 | 0.0001 |
| Colaneri et al.<sup>32</sup> | 5/21 (23.8%) | 6/21 (28.6%) | 0.781 | 0.197-3.106 | 3.41 | 0.726 |
| Rojas-Marte et al.<sup>33</sup> | 50/96 (52.1%) | 60/97 (61.9%) | 0.67 | 0.378-1.189 | 9.7 | 0.171 |
| Wadud et al.<sup>34</sup> | 17/44 (38.6%) | 26/50 (52%) | 0.581 | 0.255-1.323 | 6.9 | 0.196 |
| Ip et al.<sup>23</sup> | 62/134 (46.3%) | 231/413 (55.9%) | 0.678 | 0.459-1.003 | 12.2 | 0.0519 |
| Roumier et al.<sup>35</sup> | 3/30 (10%) | 9/30 (30%) | 0.259 | 0.0623-1.079 | 3.24 | 0.0635 |
| Guaraldi et al.<sup>36</sup> | 13/179 (7.3%) | 73/365 (20%) | 0.309 | 0.2166-0.574 | 9.11 | 0.0002 |
| Patel et al.<sup>37</sup> | 11/42 (26.2%) | 11/41 (28.6%) | 0.968 | 0.355-2.565 | 5.62 | 0.947 |
| Eimer et al.<sup>38</sup> | 4/22 (18.2%) | 7/22 (31.8%) | 0.476 | 0.116-1.94 | 3.31 | 0.301 |
| Canziani et al.<sup>39</sup> | 17/64 (26.6%) | 24/64 (37.5%) | 0.603 | 0.285-1.277 | 7.61 | 0.187 |
| Gokhale et al.<sup>40</sup> | 33/70 (47.1%) | 61/91 (67%) | 0.345 | 0.185-0.644 | 8.85 | 0.0008 |
| Rossotti et al.<sup>41</sup> | 20/74 (26.4%) | 86/146 (58.9%) | 0.517 | 0.301-0.887 | 9.23 | 0.0166 |
| Somers et al.<sup>42</sup> | 14/78 (17.9%) | 22/76 (35.5%) | 0.379 | 0.189-0.836 | 7.67 | 0.0151 |
| Potere et al.<sup>43</sup> | 2/40 (5%) | 12/40 (30%) | 0.123 | 0.0254-0.593 | 2.76 | 0.009 |
| **Total (random effects)** | **262/1008 (26.0%)** | **667/1537 (43.4%)** | **0.453** | **0.376-0.547** | **100** | **<0.001** |

Combined mortality for the TCZ-treated and SOC groups were 26.0% and 43.4% respectively. All of the studies trended toward lower mortality from severe COVID-19 with TCZ versus the SOC with the exception of Patel et al. Six studies yielded a statistically significant result (Capra et al., Guaraldi et al., Gokhale et al., Rossotti et al., Somers et al., Potere et al.) (Fig. 3). A random effects odds ratio analysis generated an odds ratio of 0.453 (95% CI 0.376-0.547) with a p-value less than 0.001. A sample size analysis with alpha of 0.01 and power of 90% determined that 392 total case and control patients are needed to detect a difference between 26.0% and 43.4% mortality. With the exception of Capra et al., the studies also scattered symmetrically around the overall odds ratio from the analysis suggesting a low likelihood of publication bias (Fig. 4). TCZ patients in five studies had significant secondary bacteremia but one reported a lower rate than the SOC (Rojas Marte et al.). Eight studies reported no adverse effects from TCZ.

**Figure 3:**
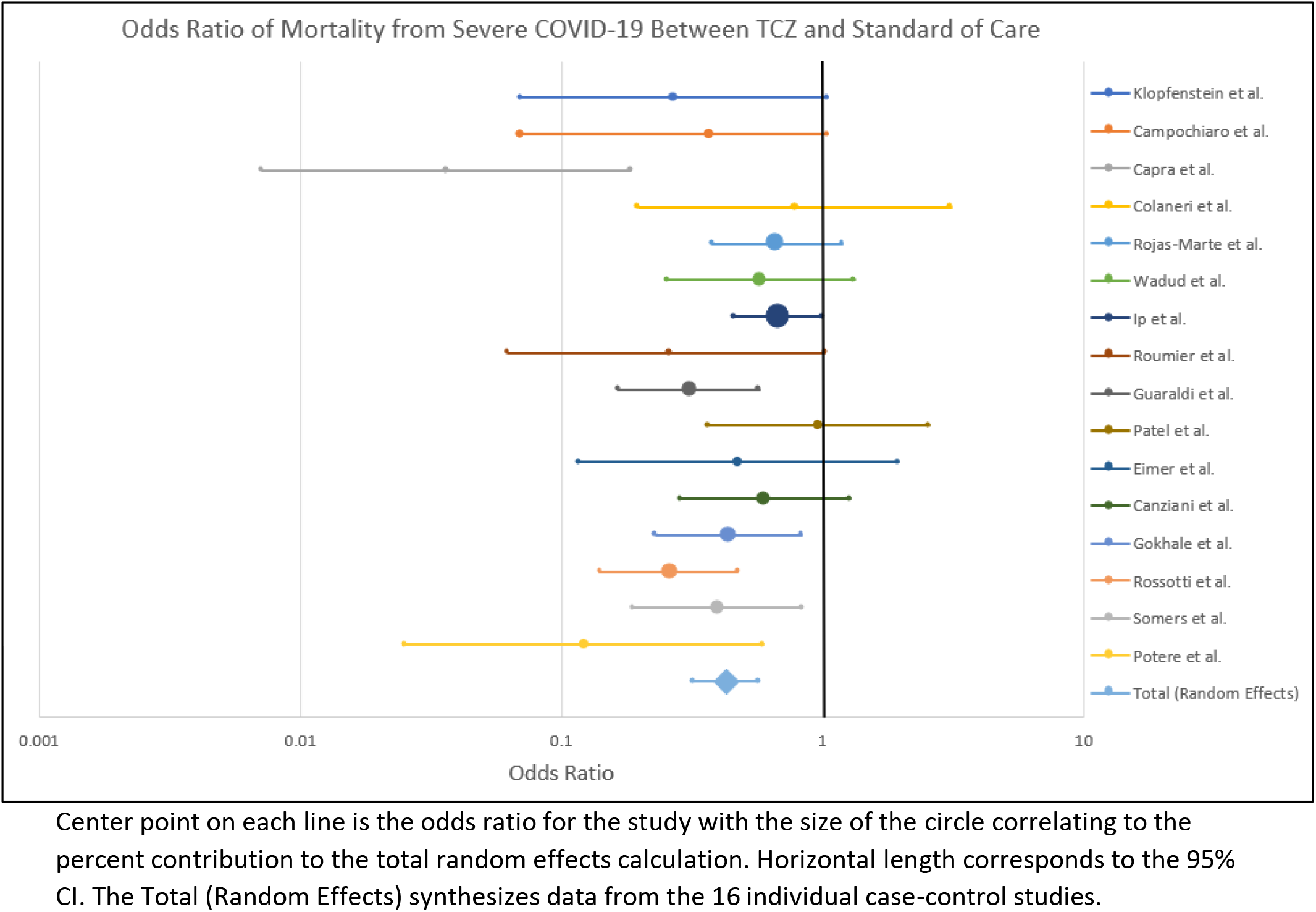
Forrest plot depicting odds ratio for death from severe COVID-19 with TCZ versus the SOC.

**Figure 4:**
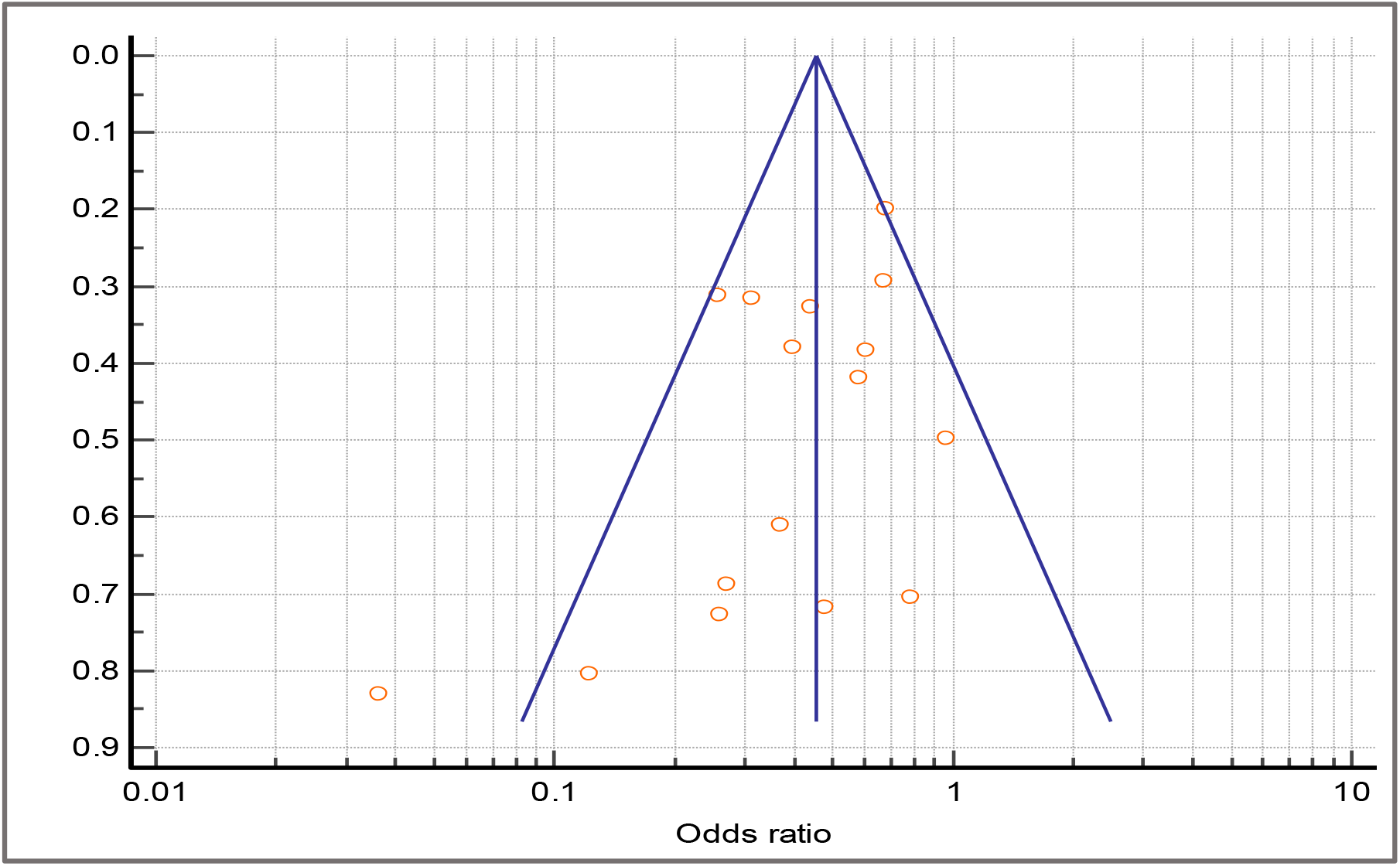
Funnel plot relating the odds ratio to the standard of the effect estimate for each study. Vertical blue line depicts the overall random effects odds ratio.

Center point on each line is the odds ratio for the study with the size of the circle correlating to the percent contribution to the total random effects calculation. Horizontal length corresponds to the 95% CI. The Total (Random Effects) synthesizes data from the 16 individual case-control studies.

Several variables varied between studies that could impact the mortality rate results. These include SOC, observation time, TCZ administration, treatment date, baseline clinical characteristics, geographic location and resources and mean/median age. Study design was also an important varying factor that may change results as nine of the sixteen studies matched cases with controls and thirteen studies were retrospective as opposed to prospective cohort.

### Uncontrolled Studies

The 18 uncontrolled trials encompassed 886 total patients who received TCZ. The mortality rate from severe COVID-19 ranged from 0% to 42.4% (SD 9.87%), although the two studies with 0% had relatively small sample sizes (20 for Xu et al. and 12 for Borku Uysal et al.). The raw overall mortality rate from the 12 studies is 16.0%. The initial patient severity level ranged from “severe” – requiring supplemental oxygen – to ICU admission. One study only investigated ICU patients (Issa et al.) and others included as many as 77.7% on MV. SOC varied more widely in the uncontrolled trials than the controlled, but hydroxychloroquine was still the most common additional drug used. Few major side effects such as bacterial/fungal infections and increased hepatic enzymes were reported.

Comparing the combined data between uncontrolled (n=18) and controlled trials (n=14) with TCZ (Table 2), excluding controlled studies with all the patients initially in the ICU, the mortality rate was 19.3% and 16.0% respectively (p=0.384).

**Table 2:** Comparison of mortality rate between controlled (n=14) and uncontrolled (n=18) studies. Mortalities and total patient values are simple tallies from each study.

|  | Mortalities | Total | Mortality Rate | SD |
| --- | --- | --- | --- | --- |
| Controlled | 150 | 778 | 19.3% | 11.9% |
| Uncontrolled | 141 | 886 | 16.0% | 9.87% |
| Difference | 3.3% |  |  |  |
| 95% CI | -10.91-4.31 |  |  |  |
| Significance Value | p=0.384 |  |  |  |

## Discussion

### Controlled Studies

The purpose of this systematic analysis was to analyze and synthesize clinical data on the efficacy of TCZ treatment against severe COVID-19. In the sixteen existing case-control trials published as of August 4, 2020, combined mortality for the TCZ-treated and SOC groups were 26.0% and 43.4% respectively. All of the studies at least trended towards reduced mortality with TCZ with one exception (Fig. 2) including six with statistical significance (Capra et al., Guaraldi et al., Gokhale et al., Rossotti et al., Somers et al., Potere et al.). After performing a quantitative synthesis, the random effects odds ratio of mortality with TCZ versus the SOC was 0.453 (95% CI 0.376-0.547, p<0.001) illustrating a reported difference in patient outcomes associated with the use of TCZ. There was no indication of publication bias except for Capra et al. (Fig. 3) and the number of patients in the meta-analysis exceeded the 392 required total case and control patients to statistically detect the difference in mortality rate. TCZ also appears to be mostly benign as few serious side effects were reported in most of the studies. As expected from an immune suppressant, the most notable adverse event was secondary infection reaching as high as 32.4% of participants (Rossotti et al.) but not at a statistically greater rate than SOC. Although the value of any single controlled clinical study does not hold definitive proof of efficacy, the consistent positive trend and statistical significance from the combined data in this analysis corroborates TCZ’s potential positive effects.

It is important to acknowledge that only one of the studies (Guaraldi et al.) randomized who received TCZ which introduces the possibility for selection bias into each study’s research methodology. Specific strengths and shortcomings for the controlled trials are outlined in Supplementary Table 2, which points towards additional methodological issues in each of the studies analyzed. Common themes include short observation timing, small sample size, difference in treatment time (later patients may benefit from better SOC), variation in disease severity within groups which can confound results, and changes to TCZ administration mid-study.

Additionally, it is possible that individual studies finding a negative effect of TCZ were not published and could not be accounted for in this systematic review. Most notably, the known public results from the COVACTA trial were not yet released in published format as of late August and were therefore not included in the quantitative synthesis. It is significant that the RCT COVACTA did not find reduced COVID-19 mortality with TCZ versus SOC, placing doubt as to the drug’s potential efficacy and the ethical basis of continuing other trials. Nonetheless, there are still multiple RCTs in progress to evaluate the efficacy of TCZ (NCT04409262; NCT04372186, NCT04356937, NCT04412772). Data from this systematic review adds merit to clinical investigations despite the initial negative results from COVACTA.

### Uncontrolled Studies

Uncontrolled trials were analyzed separately to explore trends in treatment data. Recognizing the wide variation in patient outcomes between the studies, the combined mortality rate from 12 single-arm studies using TCZ against severe COVID-19 was 16.0% (SD 9.87%). However, without clinical trials, it is difficult to determine the baseline rate COVID-19 mortality for hospitalized patients to compare with the rate calculated from the uncontrolled trials. Clinical data reviews are subject to geographic and demographic differences, such as a 5700-person evaluation in New York from April 2020 that found a 21% fatality rate for hospitalized patients which is higher than other locations and later studies.^44^ Participants must be matched to controls to eliminate bias and account for other confounding factors to draw definitive conclusions. Comparing the uncontrolled and controlled trials from this systematic review, excluding controlled trials with all the patients initially in the ICU (Ip et al. and Rojas-Marte et al.), there was no significant difference in mortality rate between the two experimental approaches (p=0.384). This observation supports the assertion that the combined reported results from the uncontrolled results from this systematic analysis are potentially accurate and corroborate the merit in evaluating TCZ efficacy. Nevertheless, the uncontrolled trials should still be evaluated with some degree of skepticism. Specific shortcomings for the individual uncontrolled trials are delineated in Supplementary Table 4.

### TCZ Treatment Timing

There is a question of timing for the IL-6 blocking treatment. All of the studies evaluated included only patients who were already in a severe disease state. Given the patterns of COVID-19 pathology and immune dysregulation, it may be logical to defer TCZ until the inflammatory phase due to the positive effects of IL-6 release in the acute infection stage which theoretically prevents SARS-CoV-2 proliferation. Given the unique, aberrant immune reaction in COVID-19, some researchers argue that the optimal time to employ targeted immune suppressants such as TCZ, in order to curtail and not enhance mortality, is when patients begin to trend towards hypoxia and inflammation.^3^ However, this timing is theoretical and must be demonstrated in a RCT.

### Limitations

There are notable limitations to this systematic analysis and qualitative synthesis of TCZ. First, only one of the studies presented randomized who received TCZ, opening the possibility for selection bias and confounding factors that cannot be accounted for statistically. This systematic analysis synthesizes data from studies with different SOCs, geographies, resources, demographics, and TCZ dosing amount, number and timing. It is not possible to conclude that TCZ is efficacious in reducing COVID-19 mortality, simply that the data trends towards a lower odds ratio for mortality with incomplete generalizability. Similarly, while patients across all of the studies were at least in severe condition, the combined data still represents individuals at various stages of COVID-19. Not all of the studies offered a longitudinal time component, so an overall hazard ratio or Kaplan-Meir survival curve cannot be produced. Additionally, many patients were still in the hospital at the end of the observation period potentially skewing the mortality rate. Importantly, in the random effects odds ratio calculation, there was no control for age, sex and baseline characteristics like individual studies were able to accomplish. As noted, the uncontrolled trials on TCZ cannot be adequately evaluated without direct comparison to a control group. Finally, it is possible that studies finding a negative effect of TCZ were not published and not accounted for in this systematic review.

## Conclusions

A systematic review of the clinical data of IL-6 inhibitor tocilizumab (TCZ) for severe COVID-19 points towards efficacy in reducing mortality from the disease. There are numerous notable methodological limitations in the studies analyzed including the lack of randomization in controlled trials and potential for an inadequate evaluation due to unpublished data. The results from this systematic analysis corroborate the logic and ethical basis for ongoing phase III RCTs on TCZ. However, use of TCZ in outside the clinical trial context is discouraged until results from these clinical trials are released.

## Data Availability

All original data from this systematic review are included in the manuscript PDF

## Declarations

* All authors have seen and approved the manuscript

** We declare no conflict of interest

*** All the data from this systematic review are presented in this manuscript

**** There was no funding provided

---

**Supplementary Table 1:** Study characteristics for the controlled trials

| Study | Methods | TCZ Administration | SOC | Length of Observation |
| --- | --- | --- | --- | --- |
| **Klopfenstein et al.** | Retrospective case-control study of hospitalized COVID-19 patients, 20 of whom received TCZ and compared with 25 SOC patients in one French hospital. | 1 or 2 doses | Hydroxychloroquine or lopinavir/ritonavir and antibiotics, some corticosteroids | Until death and/or ICU admission |
| **Campochiaro et al.** | Claimed to be the first comparison of TCZ to SOC. Retrospective cohort study of 65 patients in one Italian hospital with severe COVID-19 outside the ICU. 32 were treated with TCZ and compared outcomes with standard of care after 28 days. | 2 doses [24 hrs. apart], 400 mg | Hydroxychloroquine, lopinavir/ritonavir, ceftriaxone, azithromycin, enoxaparin | 28 days |
| **Capra et al.** | Cohort study of 62 patients treated with TCZ within 4 days post-admission compared to 23 who received only SOC at one Italian hospital. Included severe, but pre-ICU or mechanical ventilation. | 1 dose, 400 mg IV or 324 mg s.c. | Lopinavir and ritonavir | Admission to April 2, 2020 |
| **Colaneri et al.** | Retrospective, observational analysis of 21 patients treated with TCZ and matched 1:1 to patients receiving standard of care (SOC) based on propensity score. Mortality assessed after 7 days. Performed at one facility in Italy. Included patients in the ICU. | 2 doses [12 hrs. apart], 8 mg/kg (up to 800 mg), | Hydroxychloroquine, azithromycin, heparin, methylprednisone | 7 days |
| **Rojas-Marte et al.** | Retrospective case-control study in a New York medical center. Included 193 patients with mild to critical COVID-19 comparing those who received TCZ against individuals who underwent SOC therapies. | Not specified | Hydroxychloroquine, and azithromycin. Some corticosteroids, anticoagulation, remdesivir, antibiotics and vasopressors. | Not specified |
| **Wadud et al.** | Retrospective case-control study of COVID-19 patients with ARDS comparing 44 treated with TCZ and 50 controls matched on age, sex, BMI and baseline inflammatory markers. Data derived from one medical facility in New York. | Not specified | Combinations of hydroxychloroquine, azithromycin and steroids | Until death or discharge |
| **Ip et al.** | Retrospective observational case-control study of tocilizumab administration for 547 ICU patients in a 13-hospital network in New Jersey. | 1 (n=104) or 2 doses (n=20) with 400mg followed by 800 mg, 8 mg/kg or 4 mg/kg | Not specified | 30 days |
| **Roumier et al.** | Prospective case-control study of TCZ treatment in 30 patients with severe COVID-19 matched 1:1 based on age, | 1 dose, 8mg/kg | Hydroxychloroquine and azithromycin | Median 8 days |

The Efficacy of IL-6 Inhibitor Tocilizumab in Reducing Severe COVID-19 Mortality: A Systematic Review
|  | gender and disease severity. Performed in one facility in France. |  |  |  |
| --- | --- | --- | --- | --- |
| **Guaraldi et al.** | Retrospective case-control analysis of multiple medical centers in Italy. Compared 179 patients who received TCZ against 365 individuals who were only given the SOC. Patients were randomly assigned for TCZ administration. | 2 doses [12 hrs. apart], 8 mg/kg IV or 162 mg s.c. | Hydroxychloroquine, azithromycin, antiretrovirals, heparin | Until death, discharge or mechanical ventilation |
| **Patel et al.** | Retrospective cohort study using data extracted Seattle medical center comparing 42 TCZ recipients matched with 41 controls. | Not specified | Not specified | 7 days |
| **Eimer et al.** | Retrospective cohort study in a Swedish medical center of ICU patients with 29 TCZ patients and 58 controls; 22 were matched 1:1. | 8 mg/kg | Not specified | 30 days |
| **Canziani et al.** | Retrospective case-control study in two Italian hospitals of 64 TCZ patients matched 1:1 with controls | 2 doses [24 hours], 8 mg/kg | Enoxaparin, lopinavir/ritonavir, hydroxychloroquine, antibiotics, glucocorticoids | 30 days |
| **Gokhale et al.** | Retrospective cohort study in an Indian medical center contrasting 70 patients who received TCZ against 91 who did not. | 1 dose, 400 mg | Antibiotics, hydroxychloroquine, ivermectin, oseltamivir, heparin, methylprednisolone | 6-week total observation period |
| **Rossotti et al.** | Retrospective analysis in a single Italian hospital of 77 TCZ-treated patients matched 1:2 with 148 SOC | 1-2 doses [12 hrs.], 8 mg/kg | Hydroxychloroquine, remdesivir, lopinavir/ritonavir | 40 days |
| **Somers et al.** | Prospective cohort study for ventilated COVID-19 patients in a Michigan hospital contrasting 78 TCZ patients against 76 SOC. | 1 dose, 8mg/kg (within 48 hours of intubation) | Combinations of hydroxychloroquine, remdesivir, corticosteroids and ACE inhibitors | 28 days, Median 67-day follow-up |
| **Potere et al.** | Retrospective case-control study in an Italian hospital comparing 40 TCZ+SOC patients against 40 SOC matched for sex and age. | 2 doses, 324 mg | Not specified | 35 days |
| Study | Adverse Effects | Mean/Median Age TCZ | Mean/Median Age Control | TCZ Sex and Clinical Characteristics |
| **Klopfenstein et al.** | None found | 76.8 (mean) | 70.7 (mean) | 45% male; higher comorbidity index and oxygen requirements than controls. Pre- |

|  |  |  |  |  |
| --- | --- | --- | --- | --- |
|  |  |  |  | ICU upon admission. |
| **Campochiaro et al.** | Bacterial/fungal infection in 13% TCZ and 12% SOC patients | 65 (mean) | 60 (mean) | 91% male; 78% non-invasive ventilation, 22% high supplemental O2 |
| **Capra et al.** | No secondary infections reported | 63 (mean) | 70 (mean) | 73% male; oxygen requirements but not mechanical ventilation (MV) |
| **Colaneri et al.** | None found | 63.74 (median) | 62.33 (median) | 90% male; all patients in "severe" condition |
| **Rojas-Marte et al.** | Bacteremia less common in TCZ (13% versus 24%). Fever, cough and shortness of breath more common in TCZ. | 60 (mean) | 60 (mean) | 77.1% male; 6.3% moderate (nasal canula), 30.2% very severe (high-flow O2/non-rebreather), 63.5% critical (intubated) |
| **Wadud et al.** | None reported | 55.5 (mean) | 66 (age) | Sex breakdown not included; statistically higher initial inflammatory markers than controls |
| **Ip et al.** | 18 (13%) of TCZ and 44 (11%) of controls had secondary bacteremia; 12 (9%) of TCZ and 25 (6%) of controls had secondary pneumonia | 62 (mean) | 60 (mean) | 28% male; all initially in ICU |
| **Roumier et al.** | Mild hepatic cytolysis (n=2) and ventilator-acquired pneumonia (n=1) | 62.3 (mean) | 60.6 (mean) | 80% male; 23.3% in ICU at baseline |
| **Guaraldi et al.** | One patient had an injection site reaction, 1 episode of neutropenia, 1 HSV1 reactivation causing liver sepsis | 64 (mean) | 69 (mean) | 71% male; O2 support required, no MV |
| **Patel et al.** | None reported | 68 (median) | Not specified | 50% severe, 50% critical |
| **Eimer et al.** | None reported | 56 (median) | 56 (median) | ICU |
| **Canziani et al.** | Some bleeding, thrombosis and secondary infections | 63 (mean) | 64 (mean) | 73% male |
| **Gokhale et al.** | None reported | 52 (median) | 55 (median) | 67.1% male; "severe," pre ventilation |
| **Rossotti et al.** | Increased hospital stay, 32.4% infectious complications mostly in ICU, 1 death from sepsis | 59 (median) | 59 (median) | 82.4% male; severe, presence of respiratory distress |
| **Somers et al.** | Increased proportions of superinfections (56%), but no difference in 58-day mortality | 55 (mean) | 60 (mean) | 68% male; patients on MV |
| **Potere et al.** | No clinically serious adverse events, 2.5% bacterial pneumonia | 56 (mean) | 54.5 (mean) | 65% male; MV |

**Supplementary Table 2.**
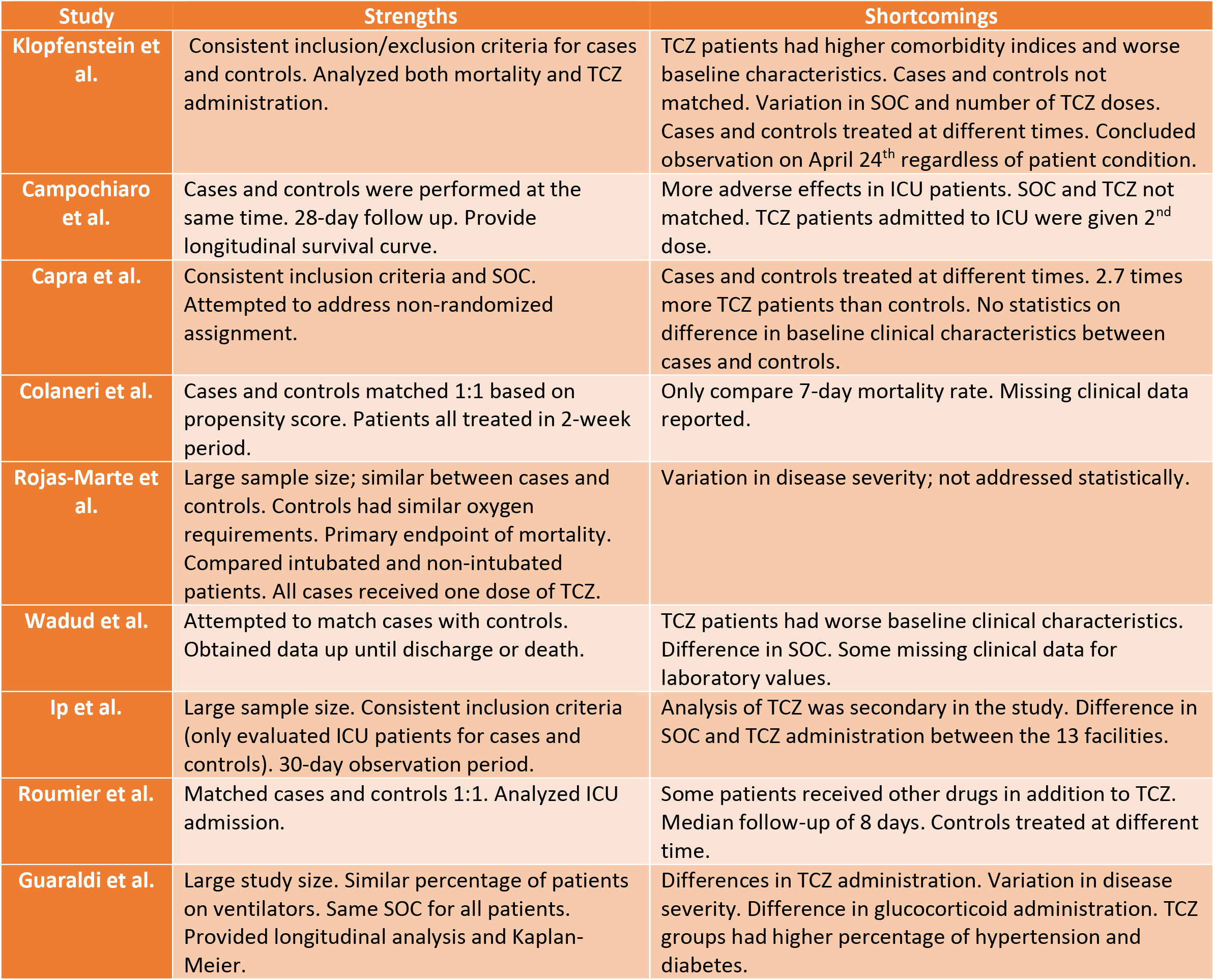
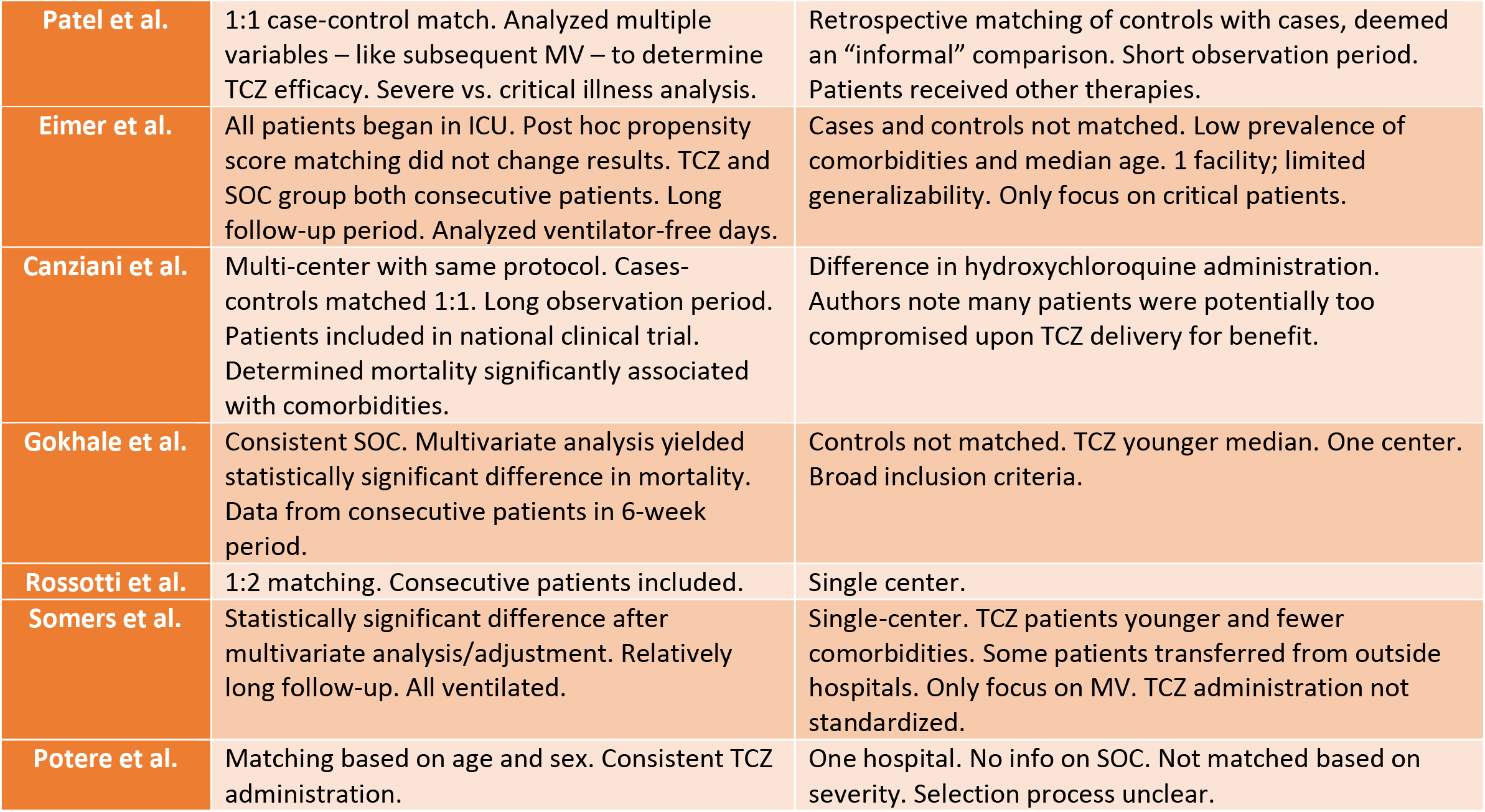
Individual subjective strengths and additional specific shortcomings of controlled trials

| Study | Strengths | Shortcomings |
| --- | --- | --- |
| **Klopfenstein et al.** | Consistent inclusion/exclusion criteria for cases and controls. Analyzed both mortality and TCZ administration. | TCZ patients had higher comorbidity indices and worse baseline characteristics. Cases and controls not matched. Variation in SOC and number of TCZ doses. Cases and controls treated at different times. Concluded observation on April 24<sup>th</sup> regardless of patient condition. |
| **Campochiaro et al.** | Cases and controls were performed at the same time. 28-day follow up. Provide longitudinal survival curve. | More adverse effects in ICU patients. SOC and TCZ not matched. TCZ patients admitted to ICU were given 2<sup>nd</sup> dose. |
| **Capra et al.** | Consistent inclusion criteria and SOC. Attempted to address non-randomized assignment. | Cases and controls treated at different times. 2.7 times more TCZ patients than controls. No statistics on difference in baseline clinical characteristics between cases and controls. |
| **Colaneri et al.** | Cases and controls matched 1:1 based on propensity score. Patients all treated in 2-week period. | Only compare 7-day mortality rate. Missing clinical data reported. |
| **Rojas-Marte et al.** | Large sample size; similar between cases and controls. Controls had similar oxygen requirements. Primary endpoint of mortality. Compared intubated and non-intubated patients. All cases received one dose of TCZ. | Variation in disease severity; not addressed statistically. |
| **Wadud et al.** | Attempted to match cases with controls. Obtained data up until discharge or death. | TCZ patients had worse baseline clinical characteristics. Difference in SOC. Some missing clinical data for laboratory values. |
| **Ip et al.** | Large sample size. Consistent inclusion criteria (only evaluated ICU patients for cases and controls). 30-day observation period. | Analysis of TCZ was secondary in the study. Difference in SOC and TCZ administration between the 13 facilities. |
| **Roumier et al.** | Matched cases and controls 1:1. Analyzed ICU admission. | Some patients received other drugs in addition to TCZ. Median follow-up of 8 days. Controls treated at different time. |
| **Guaraldi et al.** | Large study size. Similar percentage of patients on ventilators. Same SOC for all patients. Provided longitudinal analysis and Kaplan-Meier. | Differences in TCZ administration. Variation in disease severity. Difference in glucocorticoid administration. TCZ groups had higher percentage of hypertension and diabetes. |

|  |  |  |
| --- | --- | --- |
| **Patel et al.** | 1:1 case-control match. Analyzed multiple variables – like subsequent MV – to determine TCZ efficacy. Severe vs. critical illness analysis. | Retrospective matching of controls with cases, deemed an “informal” comparison. Short observation period. Patients received other therapies. |
| **Eimer et al.** | All patients began in ICU. Post hoc propensity score matching did not change results. TCZ and SOC group both consecutive patients. Long follow-up period. Analyzed ventilator-free days. | Cases and controls not matched. Low prevalence of comorbidities and median age. 1 facility; limited generalizability. Only focus on critical patients. |
| **Canziani et al.** | Multi-center with same protocol. Cases-controls matched 1:1. Long observation period. Patients included in national clinical trial. Determined mortality significantly associated with comorbidities. | Difference in hydroxychloroquine administration. Authors note many patients were potentially too compromised upon TCZ delivery for benefit. |
| **Gokhale et al.** | Consistent SOC. Multivariate analysis yielded statistically significant difference in mortality. Data from consecutive patients in 6-week period. | Controls not matched. TCZ younger median. One center. Broad inclusion criteria. |
| **Rossotti et al.** | 1:2 matching. Consecutive patients included. | Single center. |
| **Somers et al.** | Statistically significant difference after multivariate analysis/adjustment. Relatively long follow-up. All ventilated. | Single-center. TCZ patients younger and fewer comorbidities. Some patients transferred from outside hospitals. Only focus on MV. TCZ administration not standardized. |
| **Potere et al.** | Matching based on age and sex. Consistent TCZ administration. | One hospital. No info on SOC. Not matched based on severity. Selection process unclear. |

**Supplementary Table 3:**
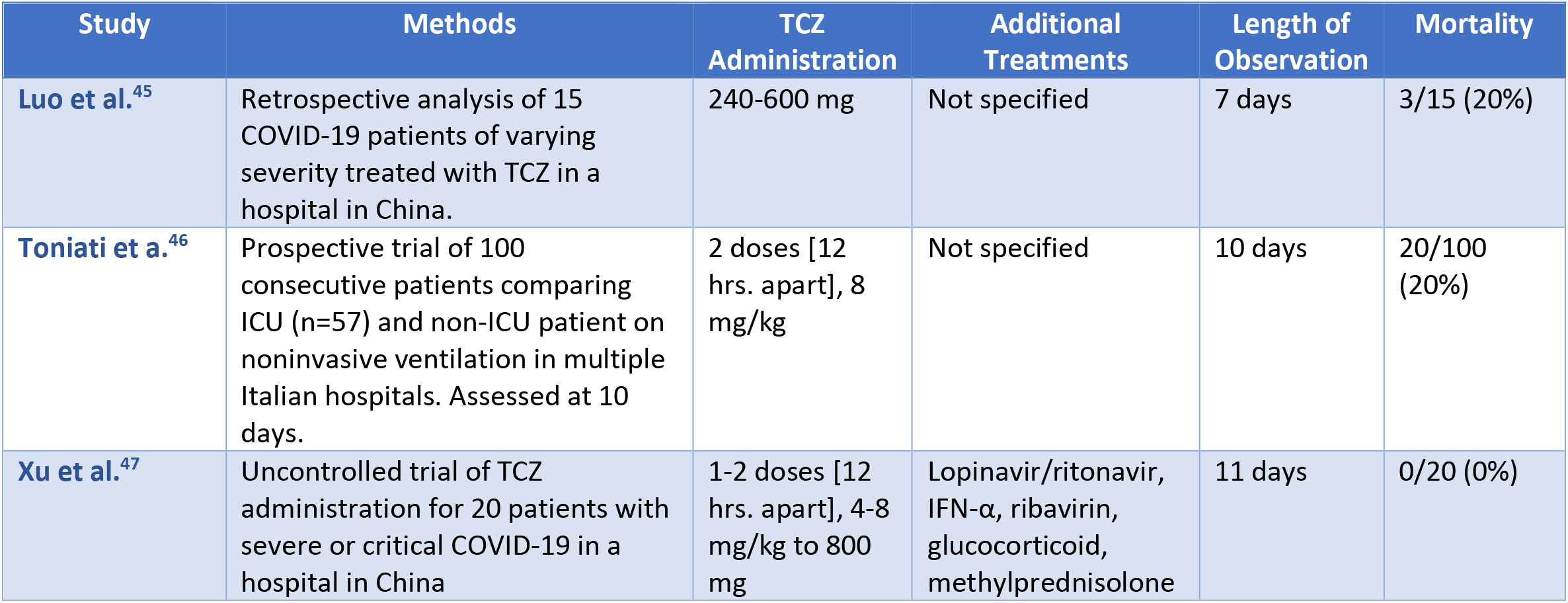
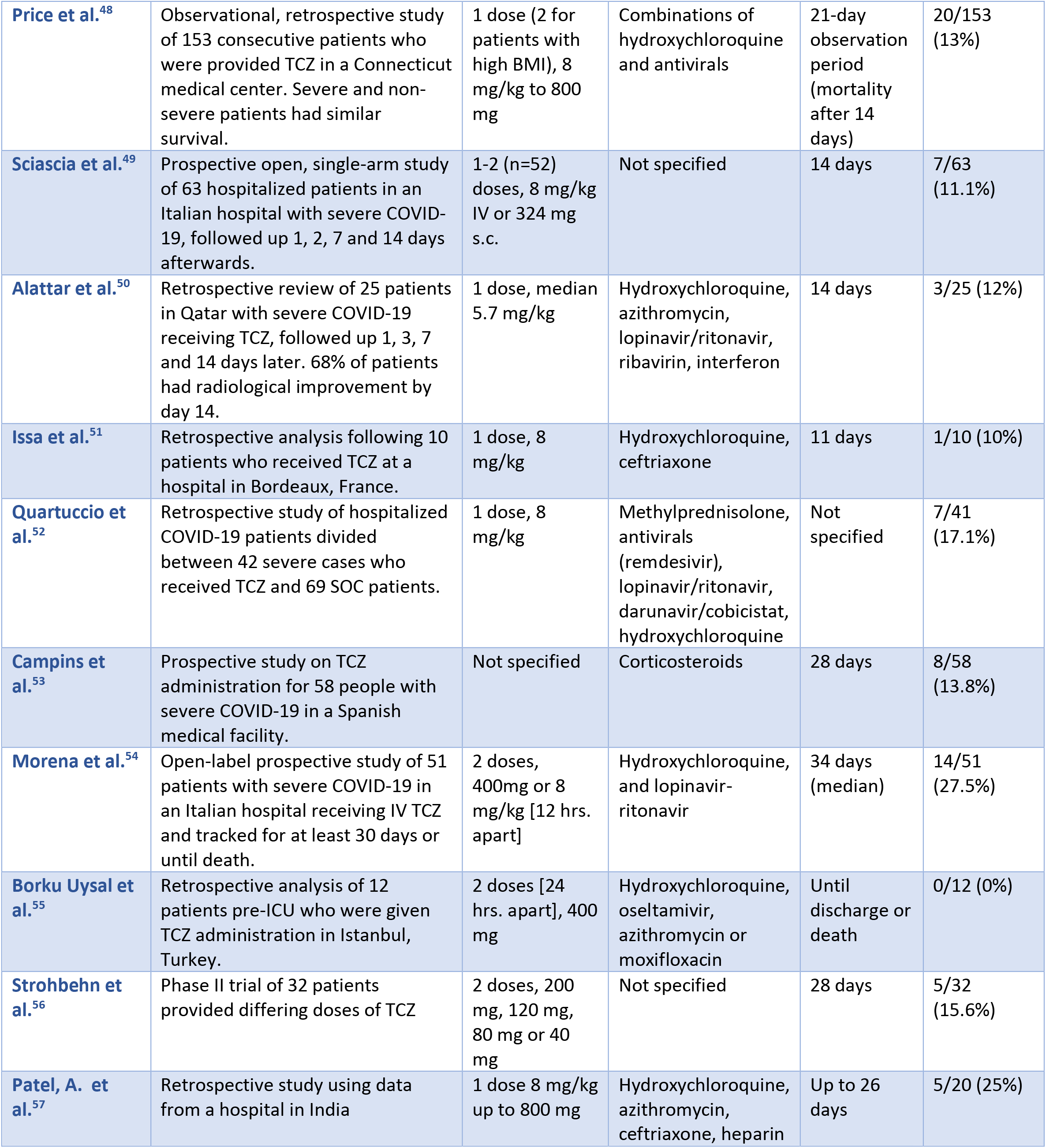
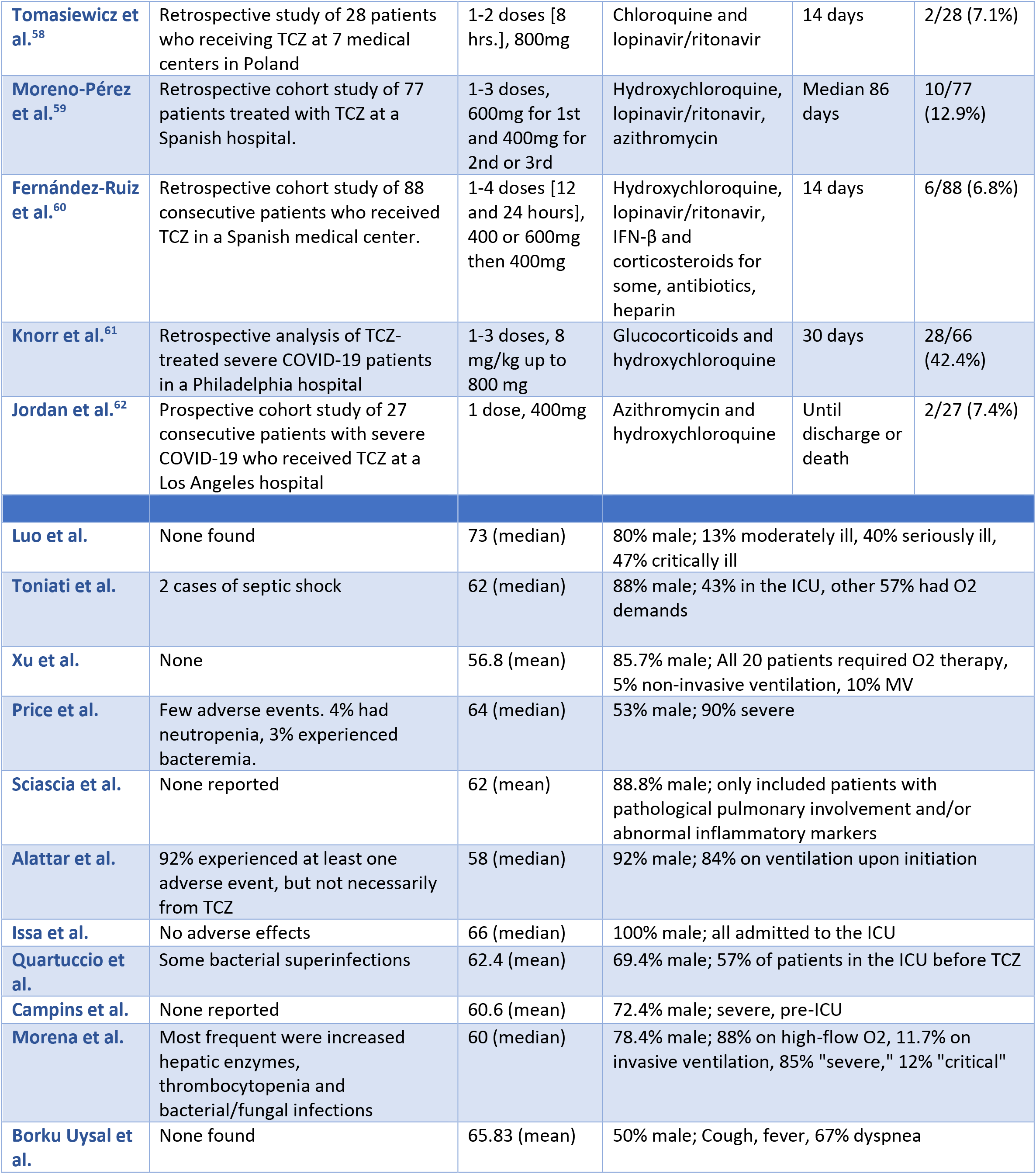
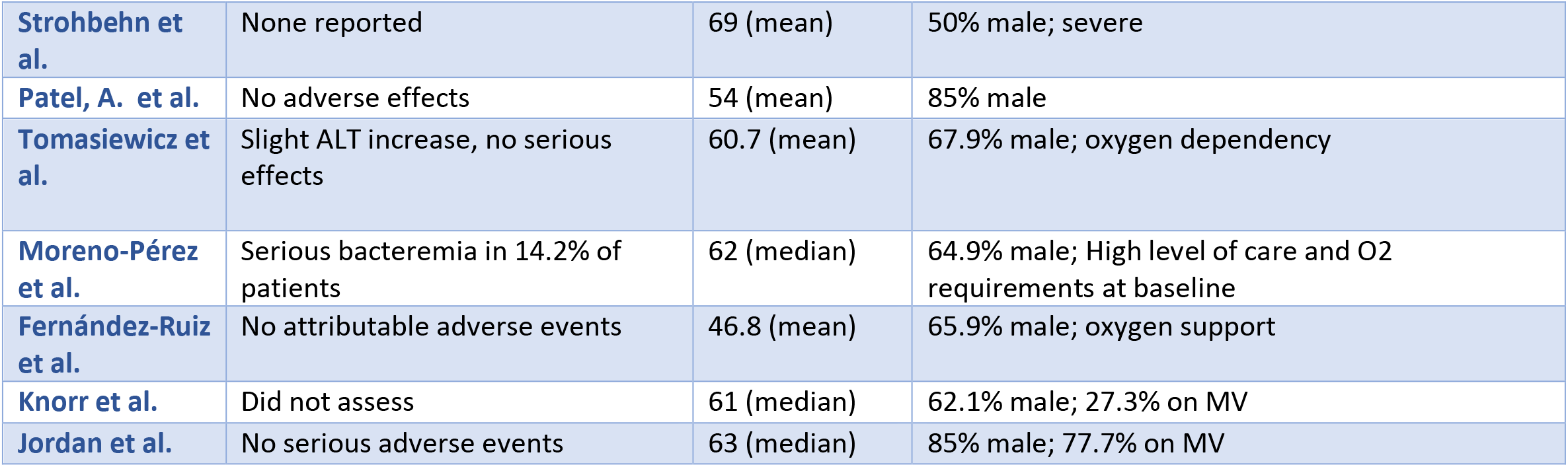
Study characteristics for the uncontrolled trials.

| Study | Methods | TCZ Administration | Additional Treatments | Length of Observation | Mortality |
| --- | --- | --- | --- | --- | --- |
| **Luo et al.<sup>45</sup>** | Retrospective analysis of 15 COVID-19 patients of varying severity treated with TCZ in a hospital in China. | 240-600 mg | Not specified | 7 days | 3/15 (20%) |
| **Toniati et al.<sup>46</sup>** | Prospective trial of 100 consecutive patients comparing ICU (n=57) and non-ICU patient on noninvasive ventilation in multiple Italian hospitals. Assessed at 10 days. | 2 doses [12 hrs. apart], 8 mg/kg | Not specified | 10 days | 20/100 (20%) |
| **Xu et al.<sup>47</sup>** | Uncontrolled trial of TCZ administration for 20 patients with severe or critical COVID-19 in a hospital in China | 1-2 doses [12 hrs. apart], 4-8 mg/kg to 800 mg | Lopinavir/ritonavir, IFN-α, ribavirin, glucocorticoid, methylprednisolone | 11 days | 0/20 (0%) |
| **Price et al.<sup>48</sup>** | Observational, retrospective study of 153 consecutive patients who were provided TCZ in a Connecticut medical center. Severe and non-severe patients had similar survival. | 1 dose (2 for patients with high BMI), 8 mg/kg to 800 mg | Combinations of hydroxychloroquine and antivirals | 21-day observation period (mortality after 14 days) | 20/153 (13%) |
| **Sciascia et al.<sup>49</sup>** | Prospective open, single-arm study of 63 hospitalized patients in an Italian hospital with severe COVID-19, followed up 1, 2, 7 and 14 days afterwards. | 1-2 (n=52) doses, 8 mg/kg IV or 324 mg s.c. | Not specified | 14 days | 7/63 (11.1%) |
| **Alattar et al.<sup>50</sup>** | Retrospective review of 25 patients in Qatar with severe COVID-19 receiving TCZ, followed up 1, 3, 7 and 14 days later. 68% of patients had radiological improvement by day 14. | 1 dose, median 5.7 mg/kg | Hydroxychloroquine, azithromycin, lopinavir/ritonavir, ribavirin, interferon | 14 days | 3/25 (12%) |
| **Issa et al.<sup>51</sup>** | Retrospective analysis following 10 patients who received TCZ at a hospital in Bordeaux, France. | 1 dose, 8 mg/kg | Hydroxychloroquine, ceftriaxone | 11 days | 1/10 (10%) |
| **Quartuccio et al.<sup>52</sup>** | Retrospective study of hospitalized COVID-19 patients divided between 42 severe cases who received TCZ and 69 SOC patients. | 1 dose, 8 mg/kg | Methylprednisolone, antivirals (remdesivir), lopinavir/ritonavir, darunavir/cobicistat, hydroxychloroquine | Not specified | 7/41 (17.1%) |
| **Campins et al.<sup>53</sup>** | Prospective study on TCZ administration for 58 people with severe COVID-19 in a Spanish medical facility. | Not specified | Corticosteroids | 28 days | 8/58 (13.8%) |
| **Morena et al.<sup>54</sup>** | Open-label prospective study of 51 patients with severe COVID-19 in an Italian hospital receiving IV TCZ and tracked for at least 30 days or until death. | 2 doses, 400mg or 8 mg/kg [12 hrs. apart] | Hydroxychloroquine, and lopinavir-ritonavir | 34 days (median) | 14/51 (27.5%) |
| **Borku Uysal et al.<sup>55</sup>** | Retrospective analysis of 12 patients pre-ICU who were given TCZ administration in Istanbul, Turkey. | 2 doses [24 hrs. apart], 400 mg | Hydroxychloroquine, oseltamivir, azithromycin or moxifloxacin | Until discharge or death | 0/12 (0%) |
| **Strohbehn et al.<sup>56</sup>** | Phase II trial of 32 patients provided differing doses of TCZ | 2 doses, 200 mg, 120 mg, 80 mg or 40 mg | Not specified | 28 days | 5/32 (15.6%) |
| **Patel, A. et al.<sup>57</sup>** | Retrospective study using data from a hospital in India | 1 dose 8 mg/kg up to 800 mg | Hydroxychloroquine, azithromycin, ceftriaxone, heparin | Up to 26 days | 5/20 (25%) |
| **Tomasiewicz et al.<sup>58</sup>** | Retrospective study of 28 patients who receiving TCZ at 7 medical centers in Poland | 1-2 doses [8 hrs.], 800mg | Chloroquine and lopinavir/ritonavir | 14 days | 2/28 (7.1%) |
| **Moreno-Pérez et al.<sup>59</sup>** | Retrospective cohort study of 77 patients treated with TCZ at a Spanish hospital. | 1-3 doses, 600mg for 1st and 400mg for 2nd or 3rd | Hydroxychloroquine, lopinavir/ritonavir, azithromycin | Median 86 days | 10/77 (12.9%) |
| **Fernández-Ruiz et al.<sup>60</sup>** | Retrospective cohort study of 88 consecutive patients who received TCZ in a Spanish medical center. | 1-4 doses [12 and 24 hours], 400 or 600mg then 400mg | Hydroxychloroquine, lopinavir/ritonavir, IFN-β and corticosteroids for some, antibiotics, heparin | 14 days | 6/88 (6.8%) |
| **Knorr et al.<sup>61</sup>** | Retrospective analysis of TCZ-treated severe COVID-19 patients in a Philadelphia hospital | 1-3 doses, 8 mg/kg up to 800 mg | Glucocorticoids and hydroxychloroquine | 30 days | 28/66 (42.4%) |
| **Jordan et al.<sup>62</sup>** | Prospective cohort study of 27 consecutive patients with severe COVID-19 who received TCZ at a Los Angeles hospital | 1 dose, 400mg | Azithromycin and hydroxychloroquine | Until discharge or death | 2/27 (7.4%) |
| **Luo et al.** | None found | 73 (median) | 80% male; 13% moderately ill, 40% seriously ill, 47% critically ill |  |  |
| **Toniati et al.** | 2 cases of septic shock | 62 (median) | 88% male; 43% in the ICU, other 57% had O2 demands |  |  |
| **Xu et al.** | None | 56.8 (mean) | 85.7% male; All 20 patients required O2 therapy, 5% non-invasive ventilation, 10% MV |  |  |
| **Price et al.** | Few adverse events. 4% had neutropenia, 3% experienced bacteremia. | 64 (median) | 53% male; 90% severe |  |  |
| **Sciascia et al.** | None reported | 62 (mean) | 88.8% male; only included patients with pathological pulmonary involvement and/or abnormal inflammatory markers |  |  |
| **Alattar et al.** | 92% experienced at least one adverse event, but not necessarily from TCZ | 58 (median) | 92% male; 84% on ventilation upon initiation |  |  |
| **Issa et al.** | No adverse effects | 66 (median) | 100% male; all admitted to the ICU |  |  |
| **Quartuccio et al.** | Some bacterial superinfections | 62.4 (mean) | 69.4% male; 57% of patients in the ICU before TCZ |  |  |
| **Campins et al.** | None reported | 60.6 (mean) | 72.4% male; severe, pre-ICU |  |  |
| **Morena et al.** | Most frequent were increased hepatic enzymes, thrombocytopenia and bacterial/fungal infections | 60 (median) | 78.4% male; 88% on high-flow O2, 11.7% on invasive ventilation, 85% "severe," 12% "critical" |  |  |
| **Borku Uysal et al.** | None found | 65.83 (mean) | 50% male; Cough, fever, 67% dyspnea |  |  |

|  |  |  |  |
| --- | --- | --- | --- |
| **Strohbehn et al.** | None reported | 69 (mean) | 50% male; severe |
| **Patel, A. et al.** | No adverse effects | 54 (mean) | 85% male |
| **Tomasiewicz et al.** | Slight ALT increase, no serious effects | 60.7 (mean) | 67.9% male; oxygen dependency |
| **Moreno-Pérez et al.** | Serious bacteremia in 14.2% of patients | 62 (median) | 64.9% male; High level of care and O2 requirements at baseline |
| **Fernández-Ruiz et al.** | No attributable adverse events | 46.8 (mean) | 65.9% male; oxygen support |
| **Knorr et al.** | Did not assess | 61 (median) | 62.1% male; 27.3% on MV |
| **Jordan et al.** | No serious adverse events | 63 (median) | 85% male; 77.7% on MV |

**Supplementary Table 4:**
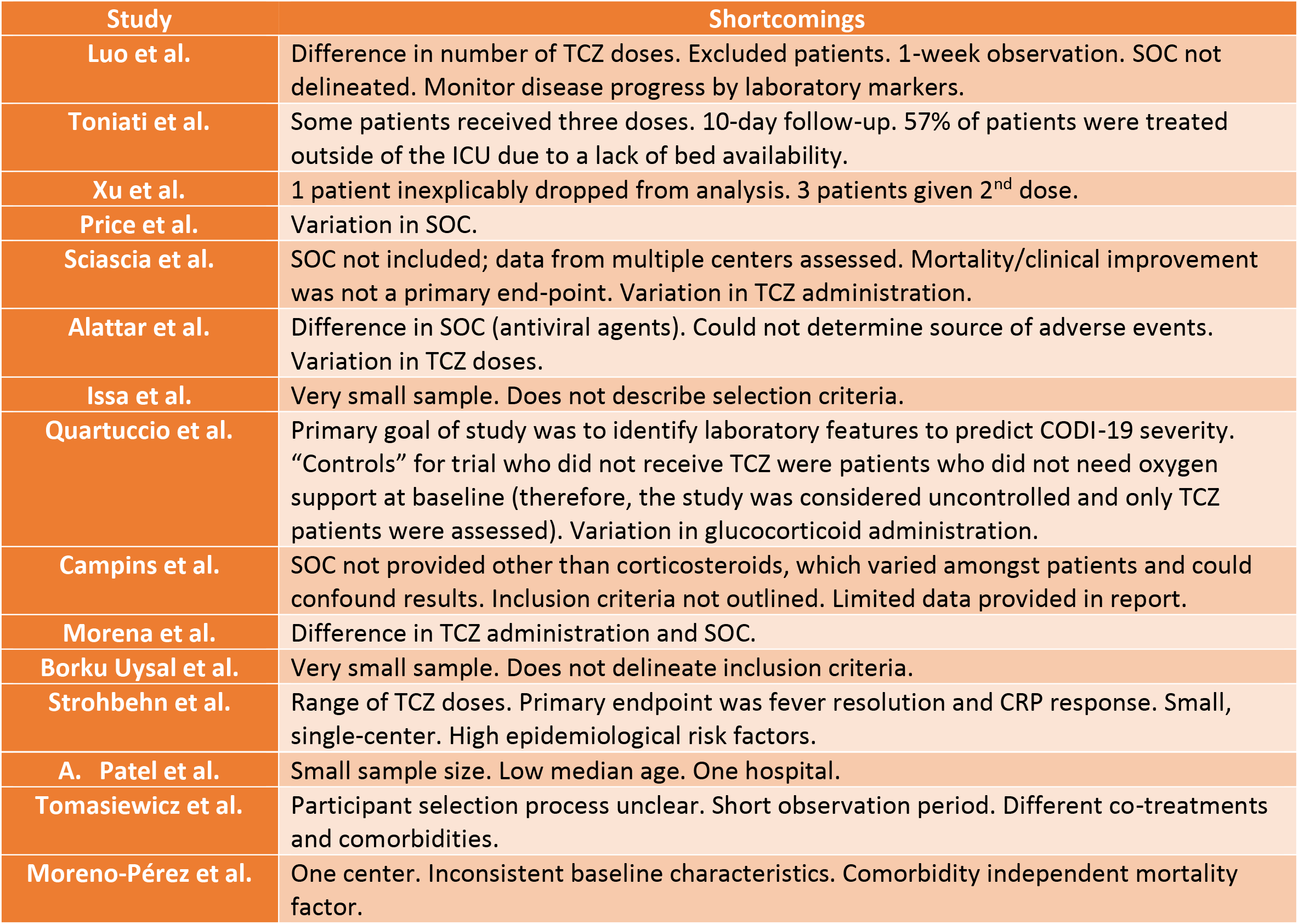
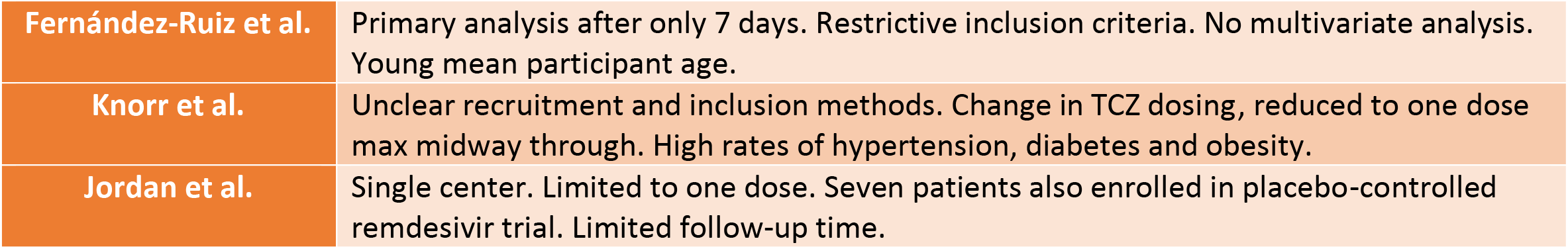
Additional specific, subjective shortcomings of uncontrolled trials

| Study | Shortcomings |
| --- | --- |
| **Luo et al.** | Difference in number of TCZ doses. Excluded patients. 1-week observation. SOC not delineated. Monitor disease progress by laboratory markers. |
| **Toniati et al.** | Some patients received three doses. 10-day follow-up. 57% of patients were treated outside of the ICU due to a lack of bed availability. |
| **Xu et al.** | 1 patient inexplicably dropped from analysis. 3 patients given 2<sup>nd</sup> dose. |
| **Price et al.** | Variation in SOC. |
| **Sciascia et al.** | SOC not included; data from multiple centers assessed. Mortality/clinical improvement was not a primary end-point. Variation in TCZ administration. |
| **Alattar et al.** | Difference in SOC (antiviral agents). Could not determine source of adverse events. Variation in TCZ doses. |
| **Issa et al.** | Very small sample. Does not describe selection criteria. |
| **Quartuccio et al.** | Primary goal of study was to identify laboratory features to predict COVID-19 severity. "Controls" for trial who did not receive TCZ were patients who did not need oxygen support at baseline (therefore, the study was considered uncontrolled and only TCZ patients were assessed). Variation in glucocorticoid administration. |
| **Campins et al.** | SOC not provided other than corticosteroids, which varied amongst patients and could confound results. Inclusion criteria not outlined. Limited data provided in report. |
| **Morena et al.** | Difference in TCZ administration and SOC. |
| **Borku Uysal et al.** | Very small sample. Does not delineate inclusion criteria. |
| **Strohbehn et al.** | Range of TCZ doses. Primary endpoint was fever resolution and CRP response. Small, single-center. High epidemiological risk factors. |
| **A. Patel et al.** | Small sample size. Low median age. One hospital. |
| **Tomasiewicz et al.** | Participant selection process unclear. Short observation period. Different co-treatments and comorbidities. |
| **Moreno-Pérez et al.** | One center. Inconsistent baseline characteristics. Comorbidity independent mortality factor. |
| **Fernández-Ruiz et al.** | Primary analysis after only 7 days. Restrictive inclusion criteria. No multivariate analysis. Young mean participant age. |
| **Knorr et al.** | Unclear recruitment and inclusion methods. Change in TCZ dosing, reduced to one dose max midway through. High rates of hypertension, diabetes and obesity. |
| **Jordan et al.** | Single center. Limited to one dose. Seven patients also enrolled in placebo-controlled remdesivir trial. Limited follow-up time. |

## Notes

### Competing Interest Statement

The authors have declared no competing interest.

### Funding Statement

No external funding was received

### Author Declarations

No IRB approval was required

### Summary of Updates

Updated Systematic Review on August 4, 2020. Updated figures and tables to match the most recent review. Addressed results from phase III trial COVACTA. Added figure to illustrate effects of IL-6.

## References

1. Huang, C., Wang, Y., Li, X. et al. Clinical features of patients infected with 2019 novel coronavirus in Wuhan, China. The Lancet. 2020; 395: 497–506

2. Docherty, A., Harrison, E., Green, C. et al. Features of 20,133 UK patients in hospital with covid-19 using the ISARIC WHO Clinical Characterization Protocol: prospective observational cohort study. BMJ. (May 22, 2020). doi:10.1136/bmj.m1985

3. Siddiqi, H. & Mehra, M. COVID-19 Illness in Native and Immunosuppressed States: A Clinical-Therapeutic Staging Proposal. Journal of Heart and Lung Transplantation. (March 25, 2020). doi:10.1016/j.healun.2020.03.012

4. Blanco-Melo, D., Nilsson-Payant, B., Liu, W. et al. Imbalance Host Response to SARS-CoV-2 Drives Development of COVID-19. Cell. (May 28, 2020). doi:10.1016/j.cell.2020.04.026

5. “Cao, X. COVID-19: immunopathology and its implications for therapy. Nature. (May 2020). doi:10.1038/s41577-020-0308-3“

6. Shi, Y. Immunopathological characteristics of coronavirus disease 2019 cases in Guangzhou, China. *medRxiv*. (March 16, 2020). doi:10.1101/2020.03.12.20034736

7. Zheng, M., Gao, Y., Wang, G. et al. Functional exhaustion of antiviral lymphocytes in COVID-19 patients. Cellular & Molecular Immunology. March 7, 2020; 17:533–535. doi:10.1038/s41423-020-0402-2

8. Jamilloux, Y., Henry, T., Belot, A. et al. Should we stimulate or suppress immune responses in COVID-19? Cytokine and anti-cytokine interventions. Autoimmunity Reviews. (April 26, 2020). doi:10.1016/j.autrev.2020.102567

9. Giamarellos-Bourboulis, E., Netea, M., Rovina, N. et al. Complex Immune Dysregulation in COVID-19 Patients with Severe Respiratory Failure. Cell Host & Microb. June 2020; 27: 1–9. doi:10.1016/j.chom.2020.04.009

10. Zhou, F., Yu, T., Du, R., et al. Clinical course and risk factors for mortality of adult inpatients with COVID-19 in Wuhan, China: a retrospective cohort study. The Lancet. (March 11, 2020);395:1054–1062. doi:10.1016/S0140-6736(20)30566-3

11. Zhang, C., Wu, Z., Li, J. et al. Cytokine release syndrome in severe COVID-19: interleukin-6 receptor antagonist tocilizumab may be the key to reduce mortality. Int J Antimicrob Agents. (May 2020). 55(5): 105954. doi:10.1016/j.ijantimicag.2020.105954

12. Gubernatorova, E., Gorshkova, E., Polinova, A. et al. IL-6: relevance for immunopathology of SARS-CoV-2. Cytokine and Growth Factor Reviews. (May 17, 2020). doi:10.1016/j.cytogfr.2020.05.009

13. Mazzoni, A., Salvati, L., Maggi, L. et al. Impaired immune cell cytotoxicity in severe COVID-19 is IL-6 dependent. J Clin Invest. (2020). doi:10.1172/JCI138554.

14. Mehta, P., McAuley, D., Brown, M., et al. COVID-19: consider cytokine storm syndromes and immunosuppression. The Lancet. (2020). doi:10.1016/S0140-6736(20)30628-0.

15. Beigel, J., Tomashek, K., Dodd, L. et al. Remdesivir for the Treatment of Covid-19 – Preliminary Report. The New England Journal of Medicine. (May 22, 2020). doi:10.1056/NEJMoa2007764

16. Hernandez, A., Roman, Y., Pasupuleti, V. et al. Hydroxychloroquine or Chloroquine for Treatment or Prophylaxis of COVID-19: A Living Systematic Review. Annals of Internal Medicine. (May 27, 2020). doi:10.7326/M20-2496

17. Russell, C., Millar, J., & Baillie J. Clinical evidence does not support corticosteroid treatment for 2019-nCoV lung injury. The Lancet. (2020). 395(10223):473–475. doi:10.1016/s0140-6736(20)30317-2.

18. Alhazzani, W., Moller, M., Arabi, Y. et al. Surviving Sepsis Campaign: guidelines on the management of critically ill adults with Coronavirus Disease 2019 (COVID-19). Intensive Care Med. 2020; 46:854–887. doi:10.1007/s00134-020-06022-5

19. Horby, P., Lim, W., Emberson, J. et al. Effect of Dexamethasone in Hospitalized Patients with COVID-19: Preliminary Report. *medRxiv*. (June 22, 2020). doi:10.1101/2020.06.22.20137273

20. “Clinical management of COVID-19.” World Health Organization. (May 27, 2020). Retrieved from https://www.who.int/publications-detail/clinical-management-of-severe-acute-respiratory-infection-when-novel-coronavirus-(ncov)-infection-is-suspected

21. Coomes E. & Haghbayan H. Interleukin-6 in COVID-19: A Systematic Review and Meta-Analysis. MedRxiv (2020). doi:10.1101/2020.03.30.20048058

22. Velazquez-Salinas, L., Verdugo-Rodriguez, A., Rodriguez, L, et al. The Role of Interleukin 6 During Viral Infections. Front Microbiol. (2019) doi:10.3389/fmicb.2019.01057.”

23. Ip, Ap., Berry, D., Hansen, E. et al. Hydroxychloroquine and Tocilizumab Therapy in COVID-19 Patients – An Observational Study. *medRxiv*. (May 25, 2020). doi:10.1101/2020.05.21.20109207

24. “Roche provides an update on the phase III COVACTA trial of Actemra/RoActemra in hospitalised patients with severe COVID-19 associated pneumonia.” Roche. (July 29, 2020). Retrieved from https://www.roche.com/investors/updates/inv-update-2020-07-29.htm

25. “Sanofi and Regeneron’s Kevzara fails in Phase III Covid-19 trial.” Clinical Trials Arena. (July 3, 2020). Retrieved from https://www.clinicaltrialsarena.com/news/kevzara-us-covid19-trial-data/

26. Alzghari, S. & Acuña, V. Supportive Treatment with Tocilizumab for COVID-19: A Systematic Review. Journal of Clinical Virology. (April 19, 2020). doi:10.1016/j.jcv.2020.104380

27. Antwi-Amoaben, D., Kanji, Z., Ford, F. et al. Clinical Outcomes in COVID-19 Patients Treated with Tocilizumab: An Individual Patient Data Systematic Review. Journal of Medical Virology. (May 2020). doi:10.1002/jmv.26038

28. Moher, D., Liberati, A., Tetzlaff, J. et al. The PRISMA Group (2009). Preferred Reporting Items for Systematic Reviews and Meta-Analyses: The PRISMA Statement. PLoS Med. (2009). 6(6). doi:10.1371/journal.pmed1000097

29. Klopfenstein, T., Zayet, S., Lohse, A. et al. Tocilizumab therapy reduced intensive care unit admissions and/or mortality in COVID-19 patients. Med Mal Infect. (May 6, 2020). doi:10.1016/j.medmal.2020.05.001

30. Campochiaro, C., Della-Torre, E., Cavalli, G. et al. Efficacy and safety of tocilizumab in severe COVID-19 patients: a single-centre retrospective cohort study. European Journal of Internal Medicine. (May 22, 2020). doi:10.1016/j.ejim.2020.05.021

31. Capra, R., Rossi, N., Mattioli, F. et al. Impact of low dose tocilizumab on mortality rate in patients with COVID-19 related pneumonia. European Journal of Internal Medicine. (May 6, 2020). doi:10.1016/j.ejim.2020.05.009

32. Colaneri, M., Bogliolo, L., Valsecchi, P. et al. Tocilizumab for Treatment of Severe COVID-19 Patients: Preliminary Results from SMAtteao COvid19 Registry (SMACORE). Microorganisms. (May 9, 2020). 10.3390/microorganisms8050695

33. Rojas-Marte, G., Khalid M., Mukhtar O. et al. Outcomes in Patients with Severe COVID-19 Disease Treated with Tocilizumab – A Case-Controlled Study. QJM. (June 23, 2020). doi:10.1093/qjmed/hcaa206

34. Wadud, N., Ahmed, N., Shergil, M. et al. Improved survival outcome in SARs-CoV-2 (COVID-19) Acute Respiratory Distress Syndrome patients with Tocilizumab administration. *medRxiv*. (May 16, 2020). doi:10.1101/2020.05.13.20100081

35. Roumier, M., Paule, R., Groh, M. et al. Interleukin-6 blockade for severe COVID-19. *medRxiv*. (March 22, 2020). doi:10.1101/2020.04.20.20061861

36. Guaraldi, G., Meschiari, M., Cozzi-Lepri, A. et al. Tocilizumab in patients with severe COVID-19: a retrospective cohort study. The Lancet. (June 24, 2020). 10.1016/S2665-9913(20)30173-9

37. Patel, K., Gooley, T., Bailey, N. et al. Use of the IL-6R Antagonist Tocilizumab in Hospitalized COVID-19 Patients. Journal of Internal Medicine, (August 4, 2020). doi:10.1111/joim.13163

38. Eimer, J., Vesterbacka, J., Svensson, A. et al. Tocilizumab shortens time on mechanical ventilation and length of hospital stay in patients with severe COVID-19: a retrospective cohort study. Journal of Internal Medicine. (August 4, 2020). doi:10.1111/joim.13162

39. Canziani, L., Trovati, S., Brunetta, E. et al. Interleukin-6 receptor blocking with intravenous tocilizumab in COVID-19 severe acute respiratory distress syndrome: A retrospective case-control survival analysis of 128 patients. Journal of Autoimmunity. (July 28, 2020). doi:10.1016/j.jaut.2020.102511

40. Gokhale, Y., Mehta, R., Karnik, N. et al. Tocilizumab improves survival in patients with persistent hypoxia in severe COVID-19 pneumonia. EClinicalMedicine. (July 22, 2020). doi:10.1016/j.eclinm.2020.100467

41. Rossotti, R., Travi, G., Ughi, N. et al. Safety and efficacy of anti-il6-receptor tocilizumab use in severe and critical patients affected by coronavirus disease 2019: A comparative analysis. Journal of Infection. (July 12, 2020). doi:10.1016/j.jinf.2020.07.008

42. Somers, E., Eschenauer, G., Troost, J. et al. Tocilizumab for treatment of mechanically ventilated patients with COVID-19. Clinical Infectious Diseases. (July 11, 2020). doi:10.1093/cid/ciaa954

43. Potere, N., Di Nisio, M., Cibelli, D. et al. Interleukin-6 receptor blockade with subcutaneous tocilizumab in severe COVID-19 pneumonia and hyperinflammation: a case-control study. Ann Rheum Dis. (July 11, 2020). doi:10.1136/annrheumdis-2020-218243

44. Richardson, S., Hirsch, J., Narasimhan, M. et al. Presenting Characteristics, Comorbidities, and Outcomes Among 5700 Patients Hospitalized With COVID-19 in the New York City Area. JAMA. (April 22, 2020); 323(20): 2052–2059. doi:10.1001/jama.2020.6775

45. Luo, P., Liu, Y., Qiu, L. et al. Tocilizumab treatment in COVID-19: A single center experience. Journal of Medical Virology. (March 26, 2020). 10.1002/jmv.25801

46. Toniati, P., Piva, S., Cattalini, M. et al. Tocilizumab for the treatment of severe COVID-19 pneumonia with hyperinflammatory syndrome and acute respiratory failure: A single center study of 100 patients in Brescia, Italy. Autoimmunity Reviews. (May 3, 2020). 19(7):102568. doi:10.1016/j.autrev.2020.102568

47. Xu, X., Han, M., Li, T. Effective treatment of severe COVID-19 patients with tocilizumab. PNAS. (May 19 2020). doi:10.1073/pnas.2005615117

48. Price, C., Altice, F., Shyr, Y. et al. Tocilizumab treatment for Cytokine Release Syndrome in hospitalized COVID-19 patients: survival and clinical outcomes. Chest. (June 20, 2020). doi:10.1016/j.chest.2020.06.006

49. Sciascia, S., Apra, F., Baffa, A. et al. Pilot prospective open, single-arm multicentre study on off-label use of tocilizumab in patients with severe COVID-19. Clinical and Experimental Rheumatology. (May 1, 2020); 38: 529–532.

50. Alattar, R., Ibrahim, T., Shaar, S. et al. Tocilizumab for the treatment of severe coronavirus disease 2019. Journal of Medical Virology. (May 5 2020). doi:10.1002/jmv.25964

51. Issa, N., Dumery, M., Guisset, O. et al. Feasibility of Tocilizumab in ICU patients with COVID-19. Journal of Medical Virology. (June 3, 2020). doi:10.1002/jmv.26110

52. Quartuccio, L., Sonaglia, A., McGonagle, D. et al. Profiling COVID-19 Pneumonia progressing into the cytokine storm syndrome: results from a single Italian Centre study on tocilizumab versus standard of care. Journal of Clinical Virology. (May 15, 2020). doi:10.1016/j.jcv.2020.104444

53. Campins, L., Boixeda, R., Perez-Cordon, L. et al. Early tocilizumab treatment could improve survival among COVID-19 patients. Clinical Experimental Rheumatology. (May 28, 2020). PMID: 32456769

54. Morena, V., Milazzo, L., Oreni, L. et al. Off-label use of tocilizumab for the treatment of SARS-CoV-2 pneumonia in Milan, Italy. European Journal of Internal Medicine. (May 21, 2020). doi:10.1016/j.ejim.2020.05.011

55. Borku Uysal, B., Ikitimur, H., Yavuzer, S., et al. Tocilizumab challenge: A series of cytokine storm therapy experience in hospitalized Covid-19 pneumonia patients. Journal of Medical Virology. (June 3, 2020). 10.1002/jmv.26111

56. Strohbehn, G., Heiss, B., Rouhani, S. et al. COVIDOSE: Low-dose tocilizumab in the treatment of Covid-19. *medRxiv*. (August 4, 2020). doi:10.1101/2020.07.20.20157503

57. Patel, A., Shah, K., Dharsandiya, M. et al. Safety and efficacy of tocilizumab in the treatment of severe acute respiratory syndrome coronavirus-2 pneumonia: A retrospective cohort study. Medical Microbiology. (July 29, 2020). doi:10.4103/ijmm.IJMM_20_298

58. Tomasiewicz, K., Piekarska, A., Stempkowska-Rejek, J. et al. Tocilizumab for patients with severe COVID-19: a retrospective, multi-center study. Expert Review of Anti-infective Therapy. (July 23, 2020). doi:10.1080/14787210.2020.1800453

59. Moreno-Pérez, O., Andres, M., Leon-Ramirez, JM. et al. Experience with tocilizumab in severe COVID-19 pneumonia after 80 days of follow-up: A retrospective cohort study. Journal of Autoimmunity. (July 22, 2020). doi:10.1016/j.jaut.2020.102523

60. Fernández‐Ruiz, M., López‐Medrano, F., Asín, M. et al.: Tocilizumab for the treatment of adult patients with severe COVID-19 pneumonia: A single-center cohort study. Journal of Medical Virology. (July 17, 2020). doi:10.1002/jmv.26308

61. Knorr J., Colomy, V., Mauriello, C. et al. Tocilizumab in patients with severe COVID-19: A single-center observational analysis. Journal of Medical Virology. (July 7, 2020). doi:10.1002/jmv.26191

62. Jordan, S., Zakowski, P., Tran, H. et al. Compassionate Use of Tocilizumab for Treatment of SARS-CoV-2. Clinical of Infectious Diseases. (June 24, 2020). doi:10.1093/cid/ciaa812

